# KL-VS heterozygosity is associated with reduced tau accumulation and lower memory impairment in Alzheimer’s disease

**DOI:** 10.1101/2020.07.29.20164434

**Authors:** Julia Neitzel, Nicolai Franzmeier, Anna Rubinski, Martin Dichgans, Matthias Brendel, Rainer Malik, Michael Ewers, for the Alzheimer’s Disease Neuroimaging Initiative (ADNI)

**Author notes:** Corresponding authors: Dr Julia Neitzel & Dr. Michael Ewers, Institute for Stroke and Dementia Research (ISD), University Hospital, LMU, Ludwig-Maximilian-University Munich, Feodor-Lynen-Straße 17, D-81377 Munich.

## Abstract

Klotho-VS heterozygosity (KL-VS^het^) is associated with reduced risk of Alzheimer’s disease (AD). However, whether KL-VS^het^ is associated with lower levels of pathologic tau, i.e. the key AD pathology driving neurodegeneration and cognitive decline, is unknown. Here, we assessed the interaction between KL-VS^het^ and levels of beta-amyloid, a key driver of tau pathology, on the levels of PET-assessed neurofibrillary tau in 354 controls and patients within the AD continuum. KL-VS^het^ showed lower cross-sectional increase in tau-PET per unit increase in amyloid-PET when compared to that of non-carriers. This effect of KL-VS^het^ on tau-PET showed a tendency to be stronger in Klotho mRNA-expressing brain regions mapped onto a gene expression atlas. KL-VS^het^ was related to better memory functions and this association was mediated by lower tau-PET. Amyloid-PET levels did not differ between KL-VS^het^ carriers versus non-carriers. Together, our findings provide evidence for a protective role of KL-VS^het^ against tau pathology and tau-related memory impairments in elderly humans at risk of AD dementia.

## INTRODUCTION

Klotho is a transmembrane protein that has been associated with enhanced longevity and better brain health in aging^1,2^. Klotho is expressed primarily in the kidney and brain, where it has been implicated in a number of vital cellular functions (for review see^3^). Loss-of-function mutations in transgenic mice are associated with reduced Klotho protein expression, accelerated aging phenotypes and dramatically shortened life span^1,4^. In humans, two variants in the KLOTHO gene (KL, 13q13.1), rs9536314 (F352V) and rs9527025 (C370S) form a functional haplotype. Carrying one copy, but not two copies of the KL-VS haplotype, referred to as KL-VS heterozygosity (KL-VS^het^), has been previously linked to increased Klotho levels in the blood^5,6^. KL-VS^het^ occurs in about 20-25% of the population^5^ and is associated with higher cognitive performance across the adult life span^5,7-9^, larger fronto-temporal grey matter volume in cognitively normal individuals^8^ and lower mortality^6,10^. Together, these results suggest a crucial role of Klotho in the maintenance of cognitive abilities and brain integrity during aging. Beyond the protective role of Klotho in normal aging, recent studies suggest an association between Klotho and reduced risk of Alzheimer’s disease (AD)^11^, the most frequent cause of dementia in the elderly^12^. A recent meta-analysis reported that KL-VS^het^ was associated with reduced AD dementia risk and cognitive decline in elderly individuals carrying the ApoE ε4 allele^13^, i.e. the strongest genetic risk factor for AD dementia possibly through elevated levels of primary AD pathology including cortical beta-amyloid (Aβ) aggregation^14,15^. Importantly, KL-VS^het^ was associated with reduced biomarker levels of Aβ deposition in ApoE ε4 carriers^16^ suggesting that KL-VS^het^ may directly alter levels of primary AD pathology.

Yet, an open question is whether Klotho is associated with altered levels of fibrillary tangles containing pathologic tau, i.e. the key driver of disease progression in AD^17^. In the presence of Aβ deposition, i.e. the earliest primary pathology in AD^18,19^, neurofibrillary tangles spread from the medial temporal lobe to higher cortical areas^20^. The progressive development of neurofibrillary tangles in the presence of Aβ pathology is closely associated with grey matter atrophy^21-23^ and cognitive worsening^20,24-26^ and is more predictive of such alterations than Aβ^27^. Due to the high clinical relevance of tau pathology, it is pivotal to understand whether the KL-VS^het^ variant attenuates the accumulation of neurofibrillary tangles at a given level of Aβ deposition, and thus cognitive decline in AD. Studies in mouse models of Aβ and accelerated aging reported that enhancing Klotho expression was associated with reduced Aβ burden and phosphorylated tau^11,28^, although conflicting results were reported as well^29^. However, these mouse models fail to develop neurofibrillary tangles in the presence of Aβ and thus only incompletely recapitulate AD-specific tau pathology in humans.

Here, we examined whether KL-VS^het^ attenuates the association between higher Aβ and higher fibrillar tau assessed via positron emission tomography (PET) in a group of 354 elderly asymptomatic and symptomatic individuals recruited within a large North American multicenter study on AD^30^. We found the KL-VS^het^ variant to be associated with an attenuated increase in regional tau-PET at pathological levels of global amyloid-PET, suggesting that KL-VS^het^ was protective against AD-related increase in neurofibrillary tangles. This effect was particularly pronounced in ApoE ε4 carriers. The strength of the KL-VS^het^ effect on region-specific tau-PET levels showed a trend to be correlated with the regional expression pattern of KL in the brain^31,32^ supporting the notion that the KL-VS^het^ variant modulates regional accumulation of tau pathology. Importantly, KL-VS^het^ was associated with higher memory performance and this effect was mediated by reduced tau-PET levels in KL-VS^het^ carriers with elevated amyloid-PET burden. For Aβ, we did not find the previously reported protective effect of KL-VS^het^ against Aβ pathology in the current sample, but confirmed this effect in a larger sample including all individuals with amyloid-PET but not necessarily tau-PET assessment available indicating that the effect size of KL-VS^het^ on Aβ was smaller than that on tau pathology. Overall, we show a protective role of KL-VS^het^ against the development of tau pathology and thus cognitive decline, suggesting that Klotho could be an attractive treatment target to slow the progression of AD.

## RESULTS

Detailed sample characteristics are presented in **Table 1**. Among the 354 participants (213 CN, 111 MCI, 30 ADD), there were 92 KL-VS^het^ carriers and 262 non-carriers. Demographics (age, sex, education) or ApoE ε4 status did not differ between KL-VS^het^ carrier and non-carrier groups (all p > 0.05). Continuous values of global amyloid-PET uptake did not differ between KL-VS^het^ carriers versus non-carriers (t(174.74) = 1.40, p = 0.163).

**Table 1.** sample characteristics.

| ADNI | KL-VS<sup>het</sup> carriers | KL-VS<sup>het</sup> non-carriers | P-value |
| --- | --- | --- | --- |
| N | 92 | 262 |  |
| Age | 71.30(6.47) | 71.21(6.26) | 0.910 |
| Sex, F:M | 50:42 | 124:138 | 0.300 |
| Diagnosis, CN:MCI:ADD | 65:21:6 | 148:90:24 | 0.057 |
| Education, y | 16.29(2.47) | 16.61(2.61) | 0.298 |
| MMSE | 27.78(2.37) | 28.38(1.96) | 0.283 |
| ApoE ε4 status, neg:pos | 59:33 | 165:97 | 0.943 |
| Global amyloid-PET, SUVR | 1.13(0.19) | 1.17(0.22) | 0.163 |

CN Cognitively Normal, MCI Mild Cognitive Impairment, ADD Alzheimer's disease dementia, F female, M male, MMSE Mini-Mental State Examination, neg: negative, pos: positive, SUVR: standard uptake value ratio

### KL-VS heterozygosity attenuates effects of amyloid on tau accumulation

In our main analysis, we tested the hypothesis that KL-VS^het^ modifies the association between Aβ and tau pathology (both assessed by continuous measures of PET uptake). In a region-of-interest (ROI) based analysis we focused on tau-PET in the inferior temporal cortex (i.e. ROI of early Aβ-related tau pathology^20,24,33-35^) and whole-brain tau-PET levels. The results of a linear regression analysis showed a significant KL-VS^het^ *x* amyloid-PET interaction effect on tau-PET levels in both the inferior temporal ROI (standardized beta = −0.57, p = 0.018, effect size measured by Cohen’s f = 0.128) and the global ROI (beta = −0.54, p = 0.030, Cohen’s f = 0.118). For both tau-PET ROIs, the increase in tau-PET as a function of rising global amyloid-PET was attenuated in KL-VS^het^ carriers versus non-carriers (**Figure 1a, b**). This result suggests that the KL-VS^het^ variant was protective against Aβ-associated accumulation of tau pathology. All analyses were controlled for main effects of the interaction terms, age, sex, diagnosis, education and ApoE ε4 carrier status. To determine whether our findings were driven by clinical diagnosis, we specifically repeated the analyses in 111 MCI patients (21 KL-VS^het^ carriers and 90 non-carriers) and found comparable KL-VS^het^ *x* amyloid-PET interaction effects on tau-PET uptake (inferior temporal ROI: beta = -1.06, p = 0.021, Cohen’s f = 0.230; global ROI: beta = -1.01, p = 0.025, Cohen’s f = 0.224; **eFigure 1 in Supplement**). Previous studies have reported an ApoE-ε4-genotype dependent effect of KL-VS^het^ on amyloid PET^16^. Hence, we additionally explored whether ApoE ε4 carriers showed a stronger protective effect of KL-VS^het^ against tau accumulation than ApoE ε4 non-carriers, controlling for age, sex, education, diagnosis and global amyloid-PET levels in the regression analyses. We found KL-VS^het^ to be associated with lower tau-PET levels in ApoE ε4 carriers for the global ROI (beta = −0.15, p = 0.032) and at trend level for the inferior temporal ROI (beta = −0.12, p = 0.080), whereas we found no main effect of KL-VS^het^ in ApoE ε4 non-carriers (global ROI: beta = 0.02, p = 0.711; inferior temporal ROI: beta=0.01, p=0.817; **eFigure 2 in Supplement**). Tested in the whole sample, the KL-VS^het^ *x* ApoE ε4 interaction effect on tau-PET levels was not significant (global ROI: beta = −0.09, p = 0.135; inferior temporal ROI: beta = −0.09, p = 0.108).

**Figure 1.**
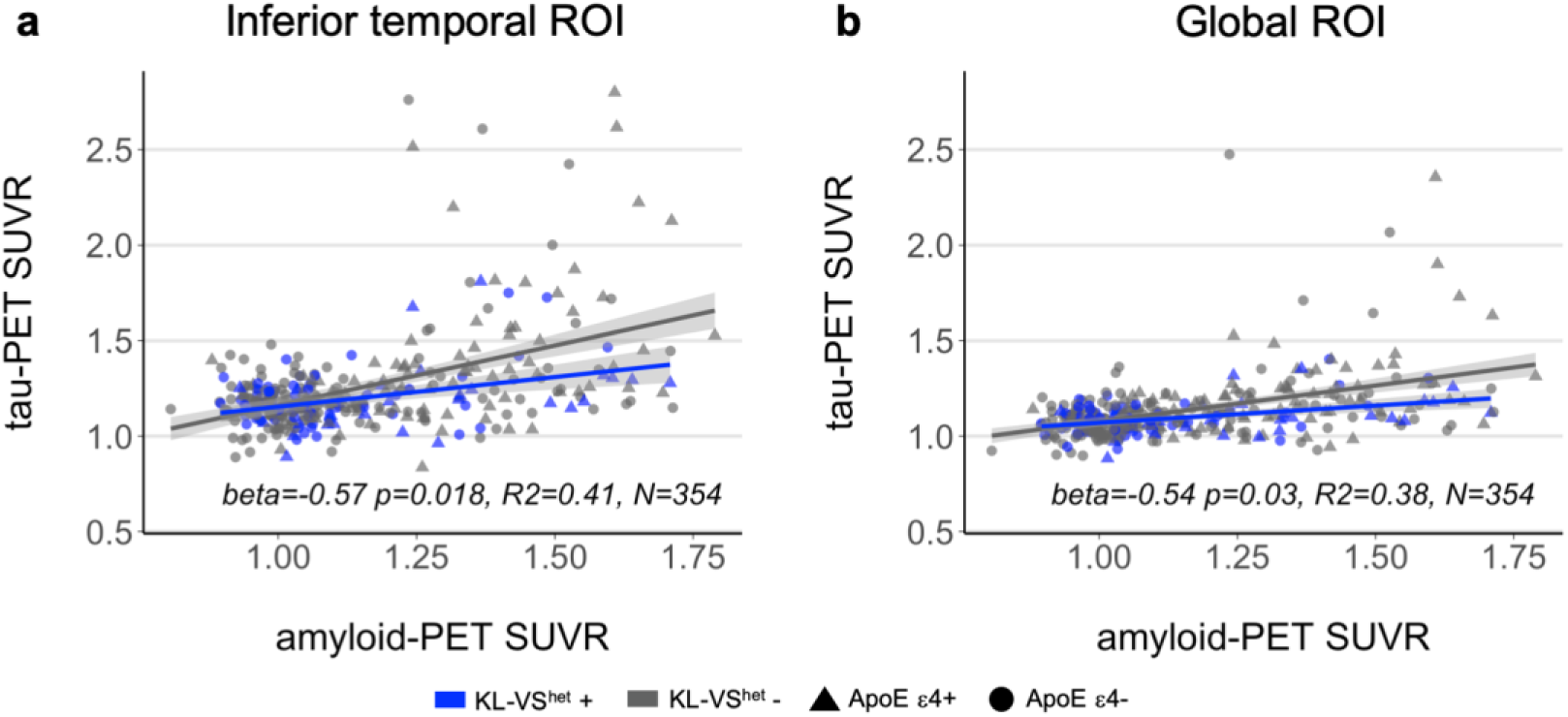
Association between KL-VS heterozygosity, amyloid- and tau-PET. Scatterplots display the relationship between global amyloid-PET levels and tau-PET-levels measured **a** in inferior temporal gyri and **b** globally in neocortical areas as a function of KL-VS^het^ variant. Blue and grey colour indicate individuals with heterozygous or non-heterozygous KL-VS alleles. Statistics of the KL-VS^het^ *x* amyloid-PET interaction effect on tau-PET uptake were derived from multiple linear regression analyses, controlled for the main effects of KL-VS^het^ and amyloid-PET levels as well as age, sex, diagnosis, education and ApoE ε4 carrier status.

**Figure 2.**
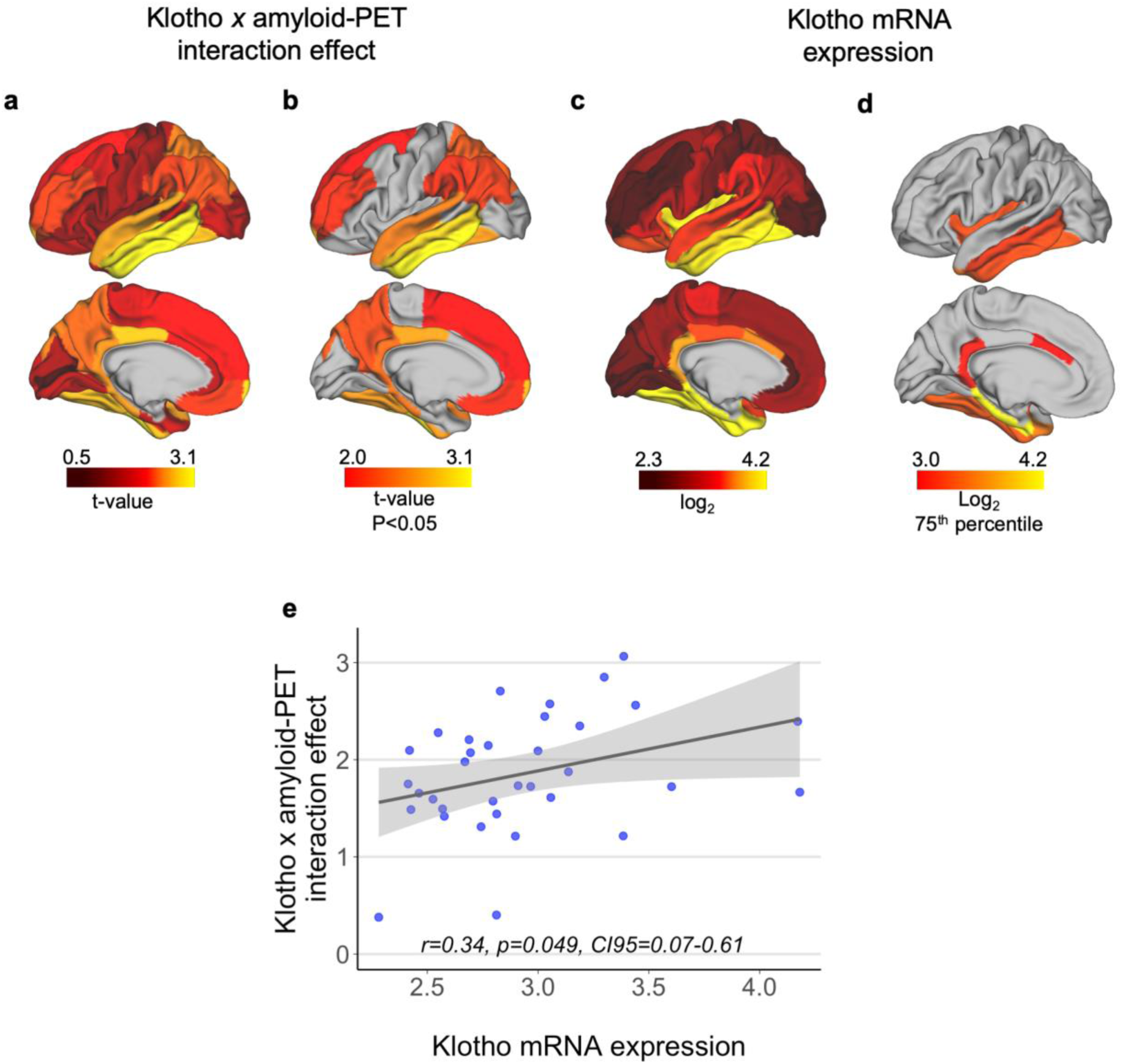
Spatial patterns of KL-VS^het^-related attenuation of tau-PET and Klotho mRNA expression. a. Surface mapping of the interaction effect between KL-VS^het^ and amyloid-PET levels on tau-PET accumulation within 34 left-hemispheric regions of the Desikan-Kiliany atlas. Yellow colours indicate higher t-values reflective of a stronger interaction effect (all t-values inverted for illustration purpose). **b** Thresholded spatial map color-code only regions with a significant (p<0.05) KL-VS^het^ *x* amyloid-PET interaction effect. **c** Surface mapping of median KL mRNA expression (i.e., log2 derived from the Allen Brain Atlas) within the identical 34 atlas regions. Yellow colours indicate higher KL mRNA expression. **d** Thresholded spatial maps restricted to regions falling above the 75th percentile of KL mRNA expression. **e** Scatterplot depicting the positive association between ROI-based KL mRNA expression and KL-VS^het^ *x* amyloid-PET interaction effect on regional tau-PET uptake. The solid grey line together with the shaded areas illustrate least-squares regression line ±95% confidence interval.

### Spatial match between KL mRNA expression and the effect of KL-VS heterozygosity on tau-PET

In order to estimate the spatial overlap between regions of KL gene expression and regions for which the KL-VS^het^ *x* amyloid-PET interaction on tau-PET was significant, we obtained whole-brain mRNA expression levels of KL generated by post-mortem microarray assessments of six healthy brain donors and subsequently mapped to the Allen Brain Atlas^31,32^. We computed median scores of log2 mRNA expression of KL across the six donors within 34 left-hemispheric regions of the Freesurfer-based Desikan–Killiany brain atlas^36^. We focused on the left hemisphere, since all donors had microarray assessment available for the left hemisphere and only two donors had assessment for the right hemisphere. Furthermore, we estimated the KL-VS^het^ *x* amyloid-PET interaction effect on tau-PET levels within the same 34 brain-atlas regions using the aforementioned regression model. Surface mapping of both the KL-VS^het^ *x* amyloid-PET interaction effect (which were all in the same direction) and KL mRNA expression is displayed in **Figure 2a-d**. Spatial correlation analysis revealed an association at trend level (r = 0.34, p = 0.049; **Figure 2e**). This result suggests that regions with higher KL mRNA expression levels were more likely to display a stronger protective effect of KL-VS^het^ on Aβ-related increase in tau-PET. Visual inspection of the thresholded spatial maps indicated that those areas showing both a significant KL-VS^het^ *x* amyloid-PET interaction effect (**Figure 2b**) and high KL mRNA expression (log2 > 75^th^ percentile) (**Figure 2d**) were specifically located within the mesio-temporal and inferior temporal brain regions and the posterior cingulum.

### Tau mediates the beneficial effect of KL-VS heterozygosity on memory

Next, we assessed whether KL-VS^het^ is beneficial for memory functions via lowering tau pathology. Because the interaction effect of KL-VS^het^ *x* amyloid-PET on tau-PET levels showed that the protective role of KL-VS^het^ against tau accumulation was stronger at higher levels of amyloid-PET, we restricted our analysis to participants who displayed elevated global amyloid-PET levels. The optimal amyloid-PET SUVR threshold for sample selection was identified in our data as SUVR > 1.0 including 67 KL-VS^het^ carriers and 191 non-carriers (see Methods for more details on sample selection). We used mediation analysis with 10,000 bootstrapping iterations in order to test whether KL-VS^het^ is associated with better memory in individuals with elevated Aβ burden and whether this effect is mediated via reduced global tau-PET levels. Memory performance was measured by an established composite score based on participant’s results across multiple different memory tests (ADNI-MEM)^37^. The mediation analysis was controlled for age, sex, education, diagnosis, ApoE ε4 status and global amyloid-PET levels. Supporting our hypothesis, we found KL-VS^het^ to be associated with higher ADNI-MEM scores (beta = 0.12, p = 0.029, Cohen’s f = 0.137, **Figure 3**) and that this relationship was significantly mediated by lower tau-PET levels (bootstrapped average causal mediation effect: beta = 0.05, 95% confidence interval = 0.01 - 0.10, p = 0.008). This result indicates that, in individuals with an elevated amyloid burden, KL-VS^het^ carriers showed less impaired episodic-memory abilities when compared to KL-VS^het^ non-carriers due to lower tau-PET levels in KL-VS^het^ carriers. A path model of the mediation analysis is shown in **Figure 4**.

**Figure 3.**
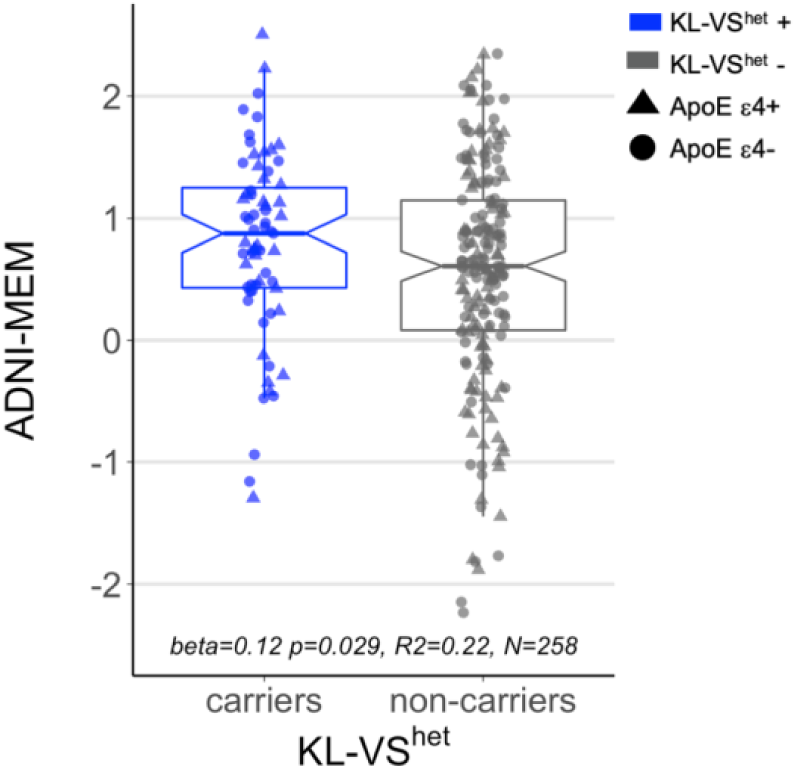
KL-VS heterozygosity effect on memory in individuals with elevated amyloid-PET burden. Boxplot shows memory performance as a function of KL-VS^het^ variant in individuals with a global amyloid-PET SUVR > 1.0. Blue and grey colour indicates individuals with heterozygous (N = 67) or non-heterozygous KL-VS alleles (N = 191). Memory was measured by an established composite score, ADNI-MEM, based on test performance across multiple different memory tests^37^. Statistical result of the main effect of KL-VS^het^ on memory was derived from multiple linear regression analysis, controlled for age, sex, diagnosis, education and ApoE ε4 carrier status.

**Figure 4.**
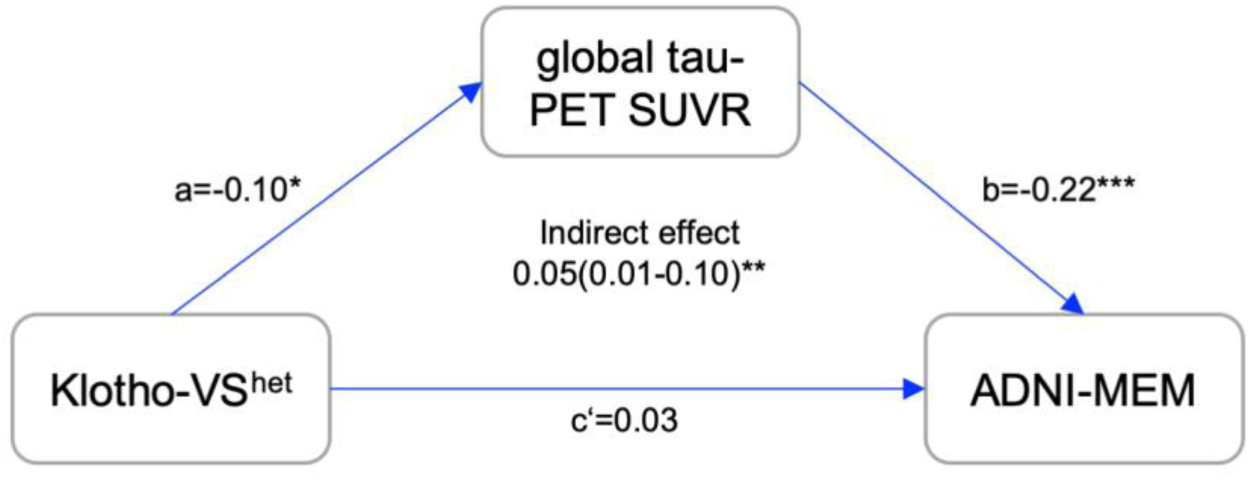
Lower Tau-PET levels mediate beneficial effect of KL-VS heterozygosity on memory in individuals with elevated amyloid-PET burden. Path diagram of the mediation model (assessed only in individuals with global amyloid-PET SUVR > 1.0, N = 258), showing that the association between KL-VS^het^ and better memory performance is mediated via lower global tau-PET uptake. Memory is measured by ADNI-MEM, i.e. an established memory composite score^37^. Path-weights are displayed as standardized beta values. All paths are controlled for age, sex, diagnosis, education, ApoE ε4 carrier status and continuous global amyloid-PET levels. Effects are reported at *p<0.05, **p<0.01 and ***p<0.001. Significance of the indirect effect was determined using bootstrapping with 10,000 iterations as implemented in the *mediation* package in *R*.

In order to address the question of whether KL-VS^het^ exerts a beneficial influence on memory via lowered neurofibrillary tau in the whole sample without stratification based on amyloid-PET levels, we estimated the interaction effect between KL-VS^het^ and amyloid-PET on ADNI-MEM scores, with and without controlling for global tau-PET levels as a covariate. We reasoned that if the association between KL-VS^het^ on cognition is mediated by tau-PET, the interaction between KL-VS^het^ and amyloid-PET on cognition should be diminished when controlling for tau-PET. Without controlling tau-PET levels, we found a borderline significant KL-VS^het^ *x* amyloid-PET interaction effect on memory functions (beta = 0.42, p = 0.049, Cohen’s f = 0.107). Specifically, we observed that individuals with high amyloid-PET burden showed better cognitive performance when being KL-VS^het^ carriers compared to non-carriers (**eFigure 3 in Supplement**). As hypothesized, the KL-VS^het^ *x* amyloid-PET interaction on memory no longer reached significance level when global tau-PET levels were controlled for (beta = 0.31, p = 0.139).

### Confirmation of the protective effect of KL-VS^het^ against Aβ accumulation in a larger sample

A recent study found a protective influence of KL-VS^het^ on longitudinal amyloid-PET in cognitively unimpaired ApoE ε4 carriers aged between 60 and 80 years, but not in ApoE ε4 non-carriers or older participants^13^. In contrast to this earlier report, we did not find an age- or ApoE ε4-dependent KL-VS^het^ effect on cross-sectional amyloid-PET levels acquired at the time of tau-PET assessment in the current sample (KL-VS^het^ *x* age interaction: beta = 0.02, p = 0.969, KL-VS^het^ *x* ApoE ε4 interaction: beta = 0.02, p = 0.744; KL-VS^het^ *x* age *x* ApoE ε4 interaction: beta = 0.39, p = 0.574; **eFigure 4 in Supplement**). However, more subtle effects may have been overlooked in the current more restricted sample of individuals undergoing both amyloid- and tau-PET. Therefore, in a supplementary analysis we included all participants with amyloid-PET (N=959) from ADNI, regardless of whether or not they underwent tau-PET assessment. We found a significant KL-VS^het^ *x* age interaction effect one amyloid-PET which demonstrated that KL-VS^het^ carriers in the lower age range (< 80 years) displayed lower amyloid-PET levels than non-carriers (beta=0.62, p=0.033, Cohen’s f = 0.069, **eFigure 5a in Supplement**). As observed earlier, the beneficial effect of KL-VS^het^ on amyloid-PET levels was driven by ApoE ε4 carriers rather than non-carriers (**eFigure 5bc in Supplement**). See **eTable 1 in Supplement** for detailed sample characteristics of the larger ADNI amyloid-PET sample compared to the current ADNI tau-PET sample. Thus, our results in the larger sample are consistent with those from Belloy et al.’s analysis on the effect of KL-VS^het^ on amyloid-PET stratified by age and ApoE genotype while also showing that the effect size of KL-VS^het^ on tau-PET is stronger than that on amyloid-PET.

## DISCUSSION

The heterozygous KLOTHO gene variant KL-VS^het^, has been previously associated with higher longevity and cognition performance in adulthood and reduced AD dementia risk^13^. We demonstrate that elderly KL-VS^het^ carriers with elevated Aβ burden, i.e. the earliest primary AD pathology, exhibited lower tau-PET levels when compared to those in KL-VS^het^ non-carriers. The KL-VS^het^ variant was associated with better memory performance, and this relationship was mediated by lower tau-PET levels, suggesting that lower levels of pathologic tau in the KL-VS^het^ carriers explain the protective association between KL-VS^het^ and memory performance. Although our findings do not implicate a causative mechanism of Klotho in AD, we provide evidence for a protective role of KL-VShet against tau pathology which is the key AD brain alteration linked to cognitive impairment.

To our knowledge, the current study is the first to date that evaluated the interaction between KL-VS^het^ and Aβ on tau accumulation and cognitive decline in humans. There is a growing literature on protective genetic variants in AD^38-40^, but only a few studies have reported genetic variants to be associated with lower tau pathology in AD^41^. For the KL-VS^het^ variant, previous studies reported an association with reduced Aβ accumulation in elderly ApoE ε4 risk-carriers^13,16^. We extend these previous findings by showing that the relationship between Aβ accumulation and fibrillar tau is modulated by KL-VS^het^, such that lower local and global tau-PET levels were observed per unit increase of global amyloid-PET burden in KL-VS^het^ carriers when compared to those in non-carriers. This is important because Aβ deposition precedes the development of dementia symptoms by up to 20 years^14^, and is associated with enhanced production of tau and the development of neurotoxic neurofibrillary tangles^42^. The region showing the strongest interaction effect between KL-VS^het^ and amyloid-PET on tau-PET was the inferior temporal gyrus (Figure 2a), a brain area that typically shows an early Aβ-related increase in tau-PET^24^ before elevated tau-PET levels extend to other higher cortical brain areas^20^. The protective association between KL-VS^het^ and tau-PET was present selectively in participants with abnormally elevated levels of amyloid-PET and in ApoE ε4 carriers. Together, these results support the notion that KL-VS^het^ is associated with an AD-related rather than age-related reduction of tau pathology.

We found KL-VS^het^ to be associated with better memory performance, mediated by the effect of KL-VS^het^ on tau-PET. Our results are broadly consistent with those from studies on healthy aging, reporting a protective effect of KL-VS^het^ on cognition^5,8-10^, and lower risk of conversion from cognitively normal to mild cognitive impairment or AD dementia in ApoE ε4 carriers^13,16^. Our findings suggest that the association between KL-VS^het^ and lower neurofibrillary tau pathology is of central importance for the protective effect of KL-VS^het^ against cognitive impairment. A previously reported absence of an association between KL-VS^het^ and cognitive decline in asymptomatic participants with elevated levels of Aβ^43^ did not assess the presence of abnormal neurofibrillary tau, which may have hampered to detect an effect of KL-VS^het^ on cognitive decline in subjects at risk of AD ^33^.

Previous studies reported KL-VS^het^ to be associated with lower amyloid-PET in ApoE ε4 carriers (but not in ApoE ε4 non-carriers), which was strongest in the age range between 60 and 80 years^13,16^. In our primary analysis we did not confirm age- or ApoE ε4-dependent effects of KL-VS^het^ on tau-PET. By investigating a larger sample of all participants with available amyloid-PET regardless of the availability of tau-PET (N = 959), we were able to substantiate those earlier findings^13,16^. Specifically, we showed reduced amyloid-PET burden in younger KL-VS^het^ carriers (< 80 years) and, in accordance with previous work, this association was mainly driven by ApoE ε4 carriers rather than non-carriers. Comparing effect sizes, Cohen’s f = 0.069 for the effect of KL-VS^het^ on amyloid-PET versus f = 0.118 for the effect on tau-PET, strengthens the important role of changes in tau pathology for understanding the role of Klotho in AD.

The mechanisms linking Klotho to tau pathology remain elusive. Klotho is a pleiotropic protein that has been implicated in multiple biological processes including insulin regulation^4^, growth factor functions, in particular of FGF23^44^, regulation of members of the redox system^45^, and calcium signaling^46^. One possibility of how the Klotho protein might be linked to reduced neurofibrillary tau is its involvement in autophagy^47^, a mechanism that is involved in the clearance of AD pathologies^48^. Lentiviral overexpression of Klotho protein in an APP-PS1 mouse model of Aβ deposition reduced Aβ plaque load in aged mice and rescued the impaired autophagy possibly by modulating the Akt/mTOR pathway^11,49^. Since APP-PS1 mice do not develop tau pathology, it remains, however, to be tested whether Klotho-induced autophagy reduces tau pathology. Those mechanistic explanations remain speculative at this point and the current work encourages future studies to investigate the mechanism that could underlie the protection Klotho exerts against the development of tau pathology.

Our findings of the spatial correspondence between the strength of the effect of KL-VS^het^ on regional tau-PET and the spatial distribution of Klotho mRNA suggests a local effect of Klotho on the development of fibrillar tau especially in temporal brain areas. The result of the spatial correlation analysis was at trend level and call for replication. Nevertheless, it is worth noting that alternative splicing of the human Klotho mRNA results in both a membrane-bound and a secreted transcript of Klotho^1,4^, indicating that Klotho may act both in a cell-autonomous manner and as a humoral factor. Therefore, differences in gene expression in Klotho in the brain and/or different circulating levels of Klotho linked to KL-VS^het5^ may influence the development of pathological tau^11^, but this link remains to be investigated.

Several caveats should be considered when interpreting the current results. Firstly, the human KL gene consists of three polymorphic variants. We decided to focus on the KL-VS haplotype given the existing evidence of its beneficial influence on Aβ and cognition in both mice and humans^5,8-10,13,50^. While the second variant C1818T (rs564481) is located on the fourth exon and likely has no functional consequences itself, the third variant G395A (rs1207568) is located in the promotor region and may be a potential regulatory site of KL. The two latter variants appear more frequent in Asian populations, where they have been linked to cardiovascular risk factors^51^. Related to the current research question, an investigation across three independent cohorts of oldest old Danes found different polymorphic variants of KL, besides KL-VS, to be associated with better cognitive functions^7^. It has yet to be proven whether these other KL variants also support resilience in AD. Another caveat is that we did not measure Klotho protein levels in the serum or CSF. Circulating levels of Klotho decrease during aging^52^ and are associated with cognitive performance^5^ and grey matter volume^53^ in cognitively unimpaired individuals. In patients with AD, CSF levels of Klotho are reduced^50^, where the experimental reversal of reduced Klotho expression in transgenic mouse models exerted beneficial effects on Aβ and cognition^8,11,28^. While the KL-VS^het^ variant has been associated with higher circulating levels of Klotho^53^, it remains to be investigated whether the association between KL-VS^het^ and pathological tau are mediated by higher protein levels in the CSF and brain tissue.

In summary, due to the high clinical relevance of tau accumulation and mixed results from trials on Aβ immunization^54^, strategies for targeting tau pathology are moving into focus^55^. Our findings revealed a protective effect of KL-VS^het^ against tau accumulation which particularly manifested in at-risk individuals, where lower tau pathology was related to better cognitive functions. Our results encourage future studies on enhancing Klotho proteins levels as a therapeutic intervention to slow down the development of tau pathology and dementia in AD.

## MATERIALS & METHODS

### Sample

354 participants were selected from ADNI phase 3 (ClinicalTrials.gov ID: NCT02854033) based on the availability of KL-VS and ApoE ε4 genotyping, T1-weighted MRI, ^18^F-AV1451 tau-PET and ^18^F-AV45 amyloid-PET. MR and PET imaging had to be acquired during the same study visit. The two single nucleotide polymorphisms (SNPs) for KL-VS (rs9536314 for F352V, rs9527025 for C370S) and ApoE (rs429358, rs7412) were genotyped using DNA extracted by Cogenics from a 3 mL aliquot of EDTA blood. Participants were assigned to the heterozygous KL-VS group when they carried 1, but not 2, copies of the KL-VS haplotype. ApoE ε4 carriers were defined as individuals carrying at least one ε4 allele. Clinical classification was performed by the ADNI centers, dividing participants into cognitively normal (CN, MMSE>24, CDR=0, non-depressed), mild cognitively impairment (MCI; MMSE>24, CDR=0.5, objective memory-loss on the education adjusted Wechsler Memory) or AD dementia (ADD; 19<MMSE<24, CDR=0.5-1.0, NINCDS/ADRDA criteria for probable AD are fulfilled). All participants provided written informed consent and all work complied with ethical regulations for work with human participants.

### MR and PET acquisition and preprocessing

All imaging data was downloaded from the ADNI loni image archive (https://ida.loni.usc.edu). Structural T1-weighted images were acquired on 3T scanners using a 3D MPRAGE sequence with 1mm isotropic voxel-size and a TR=2300ms (detailed scan protocols can be found on: https://adni.loni.usc.edu/wp-content/uploads/2017/07/ADNI3-MRI-protocols.pdf). Structural images were processed using Freesurfer (version 5.3.0) and parcellated according to the Desikan-Killiany atlas^56^.

Tau-PET was assessed in 6×5 min blocks 75 min after intravenous bolus injection of ^18^F-AV1451 Flortaucipir. Amyloid-PET scans were obtained during 4×5 min time frames measured 50-70 min post-injection of ^18^F-AV45 Florbetaben. For both AV1451 tau- and AV45 amyloid-PET we downloaded partially preprocessed data (http://adni.loni.usc.edu/methods/pet-analysis-method/pet-analysis/).

All PET images were coregistered to the corresponding T1-weighted image to make use of Freesurfer-derived masks in participants’ high resolution, native space. Standard uptake value ratio (SUVR) scores were obtained by normalizing tau-PET images to the inferior cerebellar grey matter and amyloid-PET images to the whole cerebellar grey matter, following previous recommendations^57^.

### Tau- and amyloid-PET regions of interest

For the analyses of tau-PET, we extracted mean SUVR scores from bilateral inferior temporal gyri marking amyloid-associated spread of pathology to neocortical structures ^20,24,33-35^. In addition, we assessed global tau-PET burden as the size-weighted mean SUVR score across all Freesurfer regions as described previously^58^, excluding hippocampus, thalamus and basal ganglia due to commonly reported tracer off-target binding^59^.

For the analysis of amyloid-PET images, we computed mean amyloid-PET levels from a global ROI spanning lateral and medial frontal, anterior and posterior cingulate, lateral parietal and lateral temporal regions. Mean SUVR from these regions was also used for sample stratification (see below).

### mRNA expression levels of KL

Regional gene expression was obtained from publicly available microarray measurements of regional mRNA expression based on post-mortem data from the Allen Brain Atlas (http://human.brain-map.org). The Allen Brain atlas is based on more than 60,000 microarray probes collected from 3700 autopsy-based brain tissue samples from a total of 6 individuals aged 24–57 without known history of neurological or psychiatric diseases^31,32^. Microarray-based log2 expression values of 20,737 genes within each of the 3700 samples were mapped back into MNI standard space by the Allen Brain Institute using stereotactic coordinates of the examined probes. The whole gene expression data has been recently mapped to the Freesurfer-based Desikan–Killiany atlas as median gene expression for probes falling within each of the 68 atlas ROIs^36^. Here, we specifically extracted median expression of KL mRNA within these Desikan–Killiany ROIs, to test a spatial correlation between KL expression and KL-VS^het^ effects on local tau-PET uptake. Since microarray assessments and thus KL mRNA expression of all 6 Allen brain atlas subjects were available only for the left hemisphere (vs. 2 subjects for the right hemisphere), we restricted the analysis of KL mRNA expression data to the more robust estimates of the left hemisphere in line with previous studies^60,61^.

### Neuropsychological assessment

The ADNI neuropsychological test battery contains multiple indicators for memory functions, on which basis a composite score (ADNI-MEM) has been established^37^. ADNI-MEM summarizes test performance on the Rey Auditory Verbal Learning Test, elements from the AD Assessment Scale-Cognitive Subscale, word recall from the Mini-Mental State Examination (MMSE) and from the Wechsler Logical Memory Scale II.

### Statistical Analysis

Group demographics were compared between KL-VS^het^ carriers versus non-carriers using Welch T-tests for continuous measures and Chi-squared tests for categorical measures.

In our main analysis, we tested whether KL-VS^het^ moderates the relationship between amyloid- and tau-PET. To this end, two separate multiple linear regression analyses were used to estimate the interaction effects between KL-VS^het^ and global amyloid-PET uptake. Our main analyses included tau-PET ROIs of the inferior temporal as well as global tau-PET uptake as the dependent variables. For the KL mRNA analysis (see below), we computed the interaction effect within all 34 left hemispheric regions of the Desikan-Kiliany atlas. Besides the interaction term, regression models also included the main effects of KL-VS^het^ and global amyloid-PET levels, controlled for the main effects of the interaction terms as well as potential confounding variables including age, sex, diagnosis, education and ApoE ε4 carrier status. To account for potential influences of clinical diagnoses, we ran the same analyses in the subsample of only MCI patients. Additional exploratory analyses were run to determine the influence of ApoE ε4 carrier status. In stratified analyses in either ApoE ε4 carriers or non-carriers we specifically tested for a main effect of KL-VS^het^ on tau-PET uptake including age, sex, diagnosis, education and global amyloid-PET uptake as covariates. We also examined the interaction between KL-VS^het^ and ApoE ε4 status on tau-PET levels in the whole sample taking the same covariates into account.

Next, we tested whether the favorable influence of KL-VS^het^ on local tau-PET levels overlapped within those brain regions showing higher local KL mRNA expression levels. To this end, we determined KL mRNA expression using the Allen brain atlas data in all 34 left hemispheric Desikan–Killiany atlas regions and determined the KL-VS^het^ *x* amyloid-PET interaction effect on tau-PET uptake for corresponding anatomical regions. We then tested the ROI-to-ROI Pearson–Moment correlation between regional KL mRNA expression and the interaction effect test statistic.

In addition, we tested whether KL-VS^het^ was associated with better memory functions, and whether this association was mediated by reduced tau-PET levels. Two alternative approaches were used to test this hypothesis. In the first approach, mediation analysis (causal *mediation* package implemented in *R*) was conducted in which KL-VS^het^ variant was treated as predictor, global tau-PET levels as mediator and ADNI-MEM scores as outcome. Mediation analysis was performed in the largest subsample of individuals which showed a significant main effect of KL-VS^het^ on tau-PET levels. For sample selection, we computed the main effect of KL-VS^het^ on tau-PET in different subsamples defined across a range of amyloid-PET thresholds (SUVR > 0.8-1.2). The optimal amyloid-PET threshold was identified as global SUVR > 1.0 (**eFigure 6 in Supplement**). Significance of the mediation effect was determined using 10,000 bootstrapped iterations, where each path of the model was controlled for age, sex, diagnosis, education, ApoE ε4 carrier status and global amyloid-PET levels. In an alternative approach, we estimated the interaction effect between KL-VS^het^ *x* amyloid-PET on cognitive functions. This analysis was performed in the entire sample. Importantly, the interaction analysis was once run without and once with controlling global tau-PET levels. We specifically hypothesized that if the beneficial effect of KL-VS^het^ is dependent on lowering tau accumulation, then the interaction effect should be diminished in the tau-controlled analysis. Other covariates considered in the multiple regression models were age, sex, diagnosis, education and ApoE ε4 carrier status.

Lastly, we performed an exploratory analysis with the aim to confirm previously observed age-dependent associations between KL-VS^het^ and lower amyloid-PET burden in ApoE ε4 carriers^13^. For this purpose, we tested for a KL-VS^het^ *x* age effect on global amyloid-PET levels in the current sample (N = 354) and in a larger ADNI sample (N = 959) including all participants with amyloid-PET assessment and KL-VS status (regardless of whether or not they underwent tau-PET assessment). Main effects of KL-VS^het^ and age as well as sex, education and diagnosis were considered as covariates. In sensitivity analyses, we reran the interaction model in stratified groups of either ApoE ε4 carriers or non-carriers.

All statistical analyses were conducted with *R* statistical software (version 3.6.1). P-values were considered significant when meeting a two-tailed alpha threshold of 0.05.

## Data availability

The data that support the findings of this study were obtained from the Alzheimer’s Disease Neuroimaging Initiative (ADNI) and are available from the ADNI database (adni.loni.usc.edu) upon registration and compliance with the data use agreement. A list including the anonymized participant identifiers of the currently used sample and the source file can be downloaded from the ADNI database (http://adni.loni.usc.edu/). The R-script used for the current study can be obtained from the first author upon request.

## Acknowledgement

Data used in preparation of this manuscript were obtained from the ADNI database (adni. loni.usc.edu). As such, the investigators within the ADNI study contributed to the design and implementation of ADNI and/or provided data but did not participate in analysis or writing of this paper. A complete listing of ADNI investigators can be found at: http://adni.loni.usc.edu/wp-content/uploads/how_to_apply/ADNI_Acknowledgement_List. The study was funded by DAAD post-doc fellowship (to JN), grants from the Alzheimer Forschung Initiative (AFI, Grant 15035 to ME), Legerlotz Stiftung (to ME), LMUexcellent (to ME), Hertie Foundation for Clinical Neurosciences (to NF), LMU Förderung Forschung Lehre (Reg. 1032 to NF), European Union’s Horizon 2020 research and innovation programme (grant agreement No. 666881 [SVDs@target] and 667375 [CoSTREAM]; to MD) the DFG as part of the Munich Cluster for Systems Neurology (EXC 2145 SyNergy – ID 390857198) and the CRC 1123 (B3) to MD). M.B. received speaker honoraria from GE healthcare and LMI and is an advisor of LMI. ADNI data collection and sharing for this project was funded by the ADNI (National Institutes of Health Grant U01 AG024904) and DOD ADNI (Department of Defense award number W81XWH-12-2-0012). ADNI is funded by the National Institute on Aging, the National Institute of Biomedical Imaging, and Bioengineering, and through contributions from the following: AbbVie, Alzheimer’s Association; Alzheimer’s Drug Discovery Foundation; Araclon Biotech; BioClinica, Inc.; Biogen; Bristol-Myers Squibb Company; CereSpir, Inc.; Cogstate; Eisai Inc.; Elan Phar-maceuticals, Inc.; Eli Lilly and Company; EuroImmun; F. Hoffmann-La Roche Ltd and its affiliated company Genentech, Inc.; Fujirebio; GE Healthcare; IXICO Ltd.; Janssen Alzheimer Immunotherapy Research & Development, LLC.; Johnson & Johnson Phar-maceutical Research & Development LLC.; Lumosity; Lundbeck; Merck & Co., Inc.; Meso Scale Diagnostics, LLC.; NeuroRx Research; Neurotrack Technologies; Novartis Pharmaceuticals Corporation; Pfizer Inc.; Piramal Imaging; Servier; Takeda Pharma-ceutical Company; and Transition Therapeutics. The Canadian Institutes of Health Research is providing funds to support ADNI clinical sites in Canada. Private sector contributions are facilitated by the Foundation for the National Institutes of Health (www.fnih.org).

## Contributions

J.N.: study concept and design, data processing, statistical analysis, interpretation of the results, and writing the manuscript. N.F.: critical revision of the manuscript. A.R.: data processing and critical revision of the manuscript. M.D.: critical revision of the manuscript. M.B.: critical revision of the manuscript. R.M.; .data processing and critical revision of the manuscript. M.E.: study concept and design, interpretation of the results, and writing the manuscript. ADNI provided all data used for this study.

## REFERENCES

1 Kuro-o, M. et al. Mutation of the mouse klotho gene leads to a syndrome resembling ageing. Nature 390, 45–51 (1997).

2 Zhu, Z. et al. Klotho gene polymorphisms are associated with healthy aging and longevity: Evidence from a meta-analysis. Mech. Ageing. Dev. 178, 33–40 (2019).

3 Kuro-o, M. The Klotho proteins in health and disease. Nat. Rev. Nephrol. 15, 27–44 (2019).

4 Kurosu, H. et al. Suppression of aging in mice by the hormone Klotho. Science 309, 1829–1833 (2005).

5 Dubal, D. B. et al. Life extension factor klotho enhances cognition. Cell Rep. 7, 1065–1076 (2014).

6 Arking, D. E., Atzmon, G., Arking, A., Barzilai, N. & Dietz, H. C. Association between a functional variant of the KLOTHO gene and high-density lipoprotein cholesterol, blood pressure, stroke, and longevity. Circ. Res. 96, 412–418 (2005).

7 Mengel-From, J. et al. Genetic variants in KLOTHO associate with cognitive function in the oldest old group. J. Gerontol. A Biol. Sci. 71, 1151–1159 (2016).

8 Yokoyama, J. S. et al. Variation in longevity gene KLOTHO is associated with greater cortical volumes. Ann. Clin. Transl. Neur. 2, 215–230 (2015).

9 Deary, I. J. et al. KLOTHO genotype and cognitive ability in childhood and old age in the same individuals. Neurosc. Lett. 378, 22–27 (2005).

10 de Vries, C. F. et al. Klotho, APOEε4, cognitive ability, brain size, atrophy, and survival: a study in the Aberdeen Birth Cohort of 1936. Neurobiol. Aging 55, 91–98 (2017).

11 Zeng, C.-Y. et al. Lentiviral vector–mediated overexpression of Klotho in the brain improves Alzheimer’s disease–like pathology and cognitive deficits in mice. Neurobiol. Aging 78, 18–28 (2019).

12 Barker, W. W. et al. Relative frequencies of Alzheimer disease, Lewy body, vascular and frontotemporal dementia, and hippocampal sclerosis in the State of Florida Brain Bank. Alzheimer Dis. Assoc. Disord. 16, 203–212 (2002).

13 Belloy, M. E. et al. Association of Klotho-VS Heterozygosity With Risk of Alzheimer Disease in Individuals Who Carry APOE4. JAMA Neurol. 77, 849–862 (2020).

14 Jansen, W. J. et al. Prevalence of cerebral amyloid pathology in persons without dementia: a meta-analysis. JAMA 313, 1924–1938 (2015).

15 Corder, E. H. et al. Gene dose of apolipoprotein E type 4 allele and the risk of Alzheimer’s disease in late onset families. Science 261, 921–923 (1993).

16 Erickson, C. M. et al. KLOTHO heterozygosity attenuates APOE4-related amyloid burden in preclinical AD. Neurology 92, e1878–e1889 (2019).

17 Bejanin, A. et al. Tau pathology and neurodegeneration contribute to cognitive impairment in Alzheimer’s disease. Brain 140, 3286–3300 (2017).

18 Bateman, R. J. et al. Clinical and biomarker changes in dominantly inherited Alzheimer’s disease. NEJM 367, 795–804 (2012).

19 Mattsson-Carlgren, N. et al. Abeta deposition is associated with increases in soluble and phosphorylated tau that precede a positive Tau PET in Alzheimer’s disease. Sci Adv 6, eaaz2387 (2020).

20 Schöll, M. et al. PET imaging of tau deposition in the aging human brain. Neuron 89, 971–982 (2016).

21 Harrison, T. M. et al. Longitudinal tau accumulation and atrophy in aging and alzheimer disease. Ann. Neurol. 85, 229–240 (2019).

22 LaPoint, M. R. et al. The association between tau PET and retrospective cortical thinning in clinically normal elderly. Neuroimage 157, 612–622 (2017).

23 Hanseeuw, B. J. et al. Fluorodeoxyglucose metabolism associated with tau-amyloid interaction predicts memory decline. Ann. Neurol. 81, 583–596 (2017).

24 Johnson, K. A. et al. Tau positron emission tomographic imaging in aging and early Alzheimer disease. Ann. Neurol 79, 110–119 (2016).

25 Ossenkoppele, R. et al. Associations between tau, Aβ, and cortical thickness with cognition in Alzheimer disease. Neurology 92, e601–e612 (2019).

26 Aschenbrenner, A. J., Gordon, B. A., Benzinger, T. L., Morris, J. C. & Hassenstab, J. J. Influence of tau PET, amyloid PET, and hippocampal volume on cognition in Alzheimer disease. Neurology 91, e859–e866 (2018).

27 La Joie, R. et al. Prospective longitudinal atrophy in Alzheimer’s disease correlates with the intensity and topography of baseline tau-PET. Sci Transl Med 12, eaau5732 (2020).

28 Kuang, X. et al. Klotho upregulation contributes to the neuroprotection of ligustilide in an Alzheimer’s disease mouse model. Neurobiol. Aging. 35, 169–178 (2014).

29 Dubal, D. B. et al. Life extension factor klotho prevents mortality and enhances cognition in hAPP transgenic mice. J. Neurosci. 35, 2358–2371 (2015).

30 Weiner, M. W. et al. The Alzheimer’s Disease Neuroimaging Initiative 3: Continued innovation for clinical trial improvement. Alzheimers Dement. 13, 561–571 (2017).

31 Hawrylycz, M. J. et al. An anatomically comprehensive atlas of the adult human brain transcriptome. Nature 489, 391–399 (2012).

32 Hawrylycz, M. et al. Canonical genetic signatures of the adult human brain. Nat. Neurosc. 18, 1832–1844 (2015).

33 Pontecorvo, M. J. et al. Relationships between flortaucipir PET tau binding and amyloid burden, clinical diagnosis, age and cognition. Brain 140, 748–763 (2017).

34 Lockhart, S. N. et al. Amyloid and tau PET demonstrate region-specific associations in normal older people. Neuroimage 150, 191–199 (2017).

35 Sepulcre, J. et al. In vivo tau, amyloid, and gray matter profiles in the aging brain. J. Neurosc. 36, 7364–7374 (2016).

36 French, L. & Paus, T. A FreeSurfer view of the cortical transcriptome generated from the Allen Human Brain Atlas. Front. Neurosc. 9, 323 (2015).

37 Crane, P. K. et al. Development and assessment of a composite score for memory in the Alzheimer’s Disease Neuroimaging Initiative (ADNI). Brain Imaging Behav. 6, 502–516 (2012).

38 Dumitrescu, L. et al. Genetic Variants and Functional Pathways Associated with Resilience to Alzheimer’s Disease. bioRxiv, 2020.2002.2019.954651, doi:10.1101/2020.02.19.954651 (2020).

39 Franzmeier, N. et al. The BDNF Val66Met SNP modulates the association between beta-amyloid and hippocampal disconnection in Alzheimer’s disease. Mol. Psychiatr., (2019).

40 Hohman, T. J., Dumitrescu, L., Cox, N. J. & Jefferson, A. L. Genetic resilience to amyloid related cognitive decline. Brain Imaging Behav. 11, 401–409 (2017).

41 Kleineidam, L. et al. PLCG2 protective variant p. P522R modulates Tau pathology and disease progression in patients with mild cognitive impairment. Acta Neuropathol. 139, 1025–1044 (2020).

42 Sato, C. et al. Tau Kinetics in Neurons and the Human Central Nervous System. Neuron 98, 861–864 (2018).

43 Porter, T. et al. Klotho allele status is not associated with Abeta and APOE epsilon4-related cognitive decline in preclinical Alzheimer’s disease. Neurobiol. Aging 76, 162–165 (2019).

44 Urakawa, I. et al. Klotho converts canonical FGF receptor into a specific receptor for FGF23. Nature 444, 770–774 (2006).

45 Zeldich, E. et al. The neuroprotective effect of Klotho is mediated via regulation of members of the redox system. J. Biol. 289, 24700–24715 (2014).

46 Chang, Q. et al. The b-Glucuronidase Klotho Hydrolyzes and Activates the TRPV5 Channel. Science 21, 490-493 (2005)

47 Fernandez, A. F. et al. Disruption of the beclin 1-BCL2 autophagy regulatory complex promotes longevity in mice. Nature 558, 136–140 (2018).

48 Uddin, M. S. et al. Autophagy and Alzheimer’s Disease: From Molecular Mechanisms to Therapeutic Implications. Front. Aging Neurosc.i 10, 04 (2018).

49 Kuang, X. et al. Neuroprotective effect of ligustilide through induction of α-secretase processing of both APP and Klotho in a mouse model of Alzheimer’s disease. Front. Aaging Neurosc. 9, 353 (2017).

50 Semba, R. D. et al. Klotho in the cerebrospinal fluid of adults with and without Alzheimer’s disease. Neurosc. Lett. 558, 37–40 (2014).

51 Rhee, E. et al. Relationship between polymorphisms G395A in promoter and C1818T in exon 4 of the KLOTHO gene with glucose metabolism and cardiovascular risk factors in Korean women. J. Endocrinol. Invest. 29, 613–618 (2006).

52 Yamazaki, Y. et al. Establishment of sandwich ELISA for soluble alpha-Klotho measurement: Age-dependent change of soluble alpha-Klotho levels in healthy subjects. Biochem. Biophys. Res. Commun. 398, 513–518 (2010).

53 Yokoyama, J. S. et al. Systemic klotho is associated with KLOTHO variation and predicts intrinsic cortical connectivity in healthy human aging. Brain Imaging Behav. 11, 391–400 (2017).

54 Bachurin, S. O., Bovina, E. V. & Ustyugov, A. A. Drugs in clinical trials for Alzheimer’s disease: the major trends. Medicinal research reviews 37, 1186–1225 (2017).

55 Cole, M. A. & Seabrook, G. R. On the horizon—the value and promise of the global pipeline of Alzheimer’s disease therapeutics. Alzheimers Dement. 6, e12009 (2020)..

56 Desikan, R. S. et al. An automated labeling system for subdividing the human cerebral cortex on MRI scans into gyral based regions of interest. Neuroimage 31, 968–980 (2006).

57 Landau, S. M. et al. Amyloid deposition, hypometabolism, and longitudinal cognitive decline. Ann. Neurol. 72, 578–586 (2012).

58 Maass, A. et al. Comparison of multiple tau-PET measures as biomarkers in aging and Alzheimer’s disease. Neuroimage 157, 448–463 (2017).

59 Marquié, M. et al. Validating novel tau positron emission tomography tracer [F-18]-AV-1451 (T807) on postmortem brain tissue. Ann. Neurol. 78, 787–800 (2015).

60 Grothe, M. J. et al. Molecular properties underlying regional vulnerability to Alzheimer’s disease pathology. Brain 141, 2755–2771 (2018).

61 Franzmeier, N., Rubinski, A., Neitzel, J. & Ewers, M. The BIN1 rs744373 SNP is associated with increased tau-PET levels and impaired memory. Nat. Commun. 10, 1766 (2019).

